# MR-Guided PET Denoising and Resolution Enhancement Improves Visual Interpretation and Preserves Quantitative Behavior Across Amyloid Tracers

**DOI:** 10.64898/2026.05.14.26353149

**Authors:** Caroline Szujewski, Timothy M. Shepherd, Munir Ghesani, Maria Ponisio, William Lavely, Georg Schramm, Ariane Bollack, Benjamin Aron, Gregory Lemberskiy, the Alzheimer’s Disease Neuroimaging Initiative

**Affiliations:** Microstructure Imaging, Inc., 370 Jay St FL7, Brooklyn, 11201, NY, USA; Department of Radiology, NYU Langone Health, New York, NY, USA; Department of Radiology, Mount Sinai Health System, New York, NY, USA; Mallinckrodt Institute of Radiology, Washington University in St. Louis, St. Louis, MO, USA; Northside Radiology Associates, Atlanta, GA, USA; Department of Imaging and Pathology, KU Leuven, Leuven, Belgium; GE HealthCare, Chalfont St Giles, HP8 4SP, UK; Department of Medical Physics and Biomedical Engineering, University College London, London, WC1E 6BT, UK

**Keywords:** Amyloid PET, Alzheimer’s disease, MRI-guided PET, denoising, resolution enhancement, Centiloid

## Abstract

**Background:** Amyloid-***β*** PET provides critical biomarker data for Alzheimer’s disease diagnosis and anti-amyloid therapy evaluation, yet low spatial resolution and partial volume effects result in decreased interpretability, particularly in cases with low or borderline cortical amyloid burden. While quantitative metrics (SUVr, Centiloid) aid in interpretation of amyloid burden, disagreement between visual reads and quantitative burden does occur, further blurring the line between positive or negative scans. We evaluated whether a vendor-neutral MR-guided PET denoising and resolution enhancement method (MRG) that uses Bowsher regularization improves image interpretability and reader performance while preserving established quantitative biomarkers across multiple amyloid tracers, leading to increased concordance among visual reads and quantitative metrics.

**Methods:** Standard (STN) and MRG PET images were compared for four tracers ([^**18**^F]AV-45 ([^**18**^F]florbetapir, FBP), [^**18**^F]florbetaben (FBB), [^**18**^F]flutemetamol (FMM), and [^**11**^C]Pittsburgh compound-B (PiB)) collectively from 24 MRI and 33 PET scanners. Quantitative equivalence was assessed by comparing Standardized Uptake Value ratio (SUVr) and Centiloid scores. In three of the four tracers (FBP, FBB, FMM), visual-quantitative concordance (AUC) and reader performance were evaluated in a blinded multi-reader study by four highly experienced brain PET readers who assessed image quality, artifact severity, reader confidence, and binary amyloid positivity.

**Results:** Across all tracers, MRG preserved quantitative SUVr and Centiloid metrics relative to STN (***R***^**2**^ > **0.90** for all tracers) without introducing bias to the SUVr metric. Concordance between visual reads and quantitative burden measures significantly improved with MRG. In the multi-reader study, MRG resulted in significantly higher image quality, lower artifact burden, and greater reader confidence compared to STN (***p*** < **0.0001**). Reader accuracy increased from 0.89 to 0.94, and the false-negative rate decreased from 0.08 to 0.04. Crucially, improvements in reader confidence, accuracy, and the reduction in false negative reads were most pronounced in cases with low amyloid burden near the threshold of visual positivity.

**Conclusions:** MRG denoising and resolution enhancement improved perceived image quality, reader confidence, and accuracy for amyloid PET while preserving standard quantitative behavior across tracers. By improving cortical definition in visually challenging low-burden cases without disrupting established SUVr/Centiloid behavior, MRG may reduce visual-quantitative discordance and support more confident amyloid PET interpretation near the threshold of positivity.

## 1 Introduction

Alzheimer’s disease (AD) is the most common form of dementia, currently affecting an estimated 7.2 million Americans age 65 and older, a number projected to nearly double to 13.8 million by 2060 [1]. Amyloid-*β* (A*β*) PET is central to AD diagnosis and staging, and its clinical role is expanding with the availability of FDA-approved anti-amyloid disease-modifying therapies [2–6]. As these treatments become more widely used, accurate interpretation of amyloid PET scans has become increasingly important for identifying eligible patients [7–10]. Quantitative biomarkers, including the standardized uptake value ratio (SUVr), Centiloid metric, and z-scores support A*β* PET interpretation by improving diagnostic consistency, reducing inter-rater variability, and providing an objective baseline for longitudinal monitoring [10–12].

A*β* PET is hardest to interpret for cases that have low, borderline cortical amyloid burden [10, 13, 14]. Low cortical conspicuity in these cases is further reduced by partial volume effects at the gray-white matter boundary [15–17]. Cerebral atrophy, often present in neurodegenerative disease, can further reduce cortical conspicuity and obscure delineation of the gray-white matter boundary that is essential for visual interpretation [14, 18, 19]. In clinical practice, these limitations may be compounded by aging PET systems, as longitudinal performance studies have reported progressive losses in scanner sensitivity over time, including an approximately 4.7% annual decline and a 41% reduction over 10 years before overhaul in one monitored system. Decreased PET detection sensitivity can be compensated for by increasing radiotracer dose, lengthening scan times, or accepting noisier images [20, 21]. These imagequality deficits translate directly into clinical ambiguity, especially in cases near the threshold between visually negative and visually positive scans [8, 10, 19, 22]. Although Centiloid and related quantitative metrics improve standardization and provide an objective framework for interpretation [11, 12], disagreement between quantitative burden measures and reader interpretation still occurs in clinical practice [8, 10, 12, 19]. As a result, high-quality visual reads by trained readers remain essential especially in the low-burden cases where interpretation is most fragile.

To improve amyloid PET image quality, several strategies have been proposed, including MR-informed partial volume correction and higher-performance PET systems [14, 15]. While these efforts demonstrate that cortical definition can be improved, practical barriers prevent widespread clinical adoption. Partial volume correction (PVC) can improve spatial specificity, but also amplifies noise [14, 15, 23]. At the hardware level, newer high-sensitivity PET systems demonstrate the image-quality gains that can be achieved with improved PET resolution, but these approaches depend on specialized instrumentation that may not be broadly available and may be cost-prohibitive in current routine clinical practice [24]. These efforts highlight the need for practical methods that improve cortical definition while preserving established quantitative biomarker behavior across tracers and imaging settings [13, 14]. This is particularly important in amyloid PET, where limited image contrast has historically motivated standardization approaches such as the Centiloid scale, yet challenges in visual interpretation continue to persist [10, 14, 18, 25].

In this study, we evaluated an MRI-guided (MRG) PET denoising and resolution enhancement method based on Bowsher regularization, an anatomically informed approach designed to improve structural definition while preserving quantitative PET information [26, 27]. By using structural MRI to guide PET enhancement, this approach sharpens the gray-white matter boundary, where standard amyloid PET is often most difficult to interpret [28, 29]. Structural MRI is commonly available in patients undergoing dementia evaluation, making its use for PET enhancement a practical opportunity [2, 30]. The method was implemented as a vendor-neutral post-reconstruction optimization workflow using PET DICOM inputs, supporting application across multiple scanners, tracers, and imaging settings. We tested whether MR-guided PET denoising and resolution enhancement improves image interpretability, reader confidence, and visual-quantitative concordance while preserving established quantitative biomarker behavior relative to standard reconstruction across multiple amyloid tracers and image settings.

## 2 Methods

### 2.1 Study design and cohorts

A quantitative analysis and a blinded multi-reader evaluation were performed to assess whether MRG preserves established amyloid PET quantitative behavior, improves qualitative image interpretability and reader performance, and strengthens visual-quantitative concordance relative to Standard-of-care PET (STN). Quantitative analyses used subjects from the Global Alzheimer’s Association Interactive Network (GAAIN) Centiloid Project reference data to assess preservation of established amyloid PET measures (Fig. 3). The reader study used subjects assembled from GAAIN, ADNI, and GE HealthCare clinical studies [31, 32] and was performed by four board-certified neuroradiologists to assess qualitative image interpretability, reader performance, and visual–quantitative concordance for MRG relative to STN (Figs. 4–6).

**Fig. 1:**
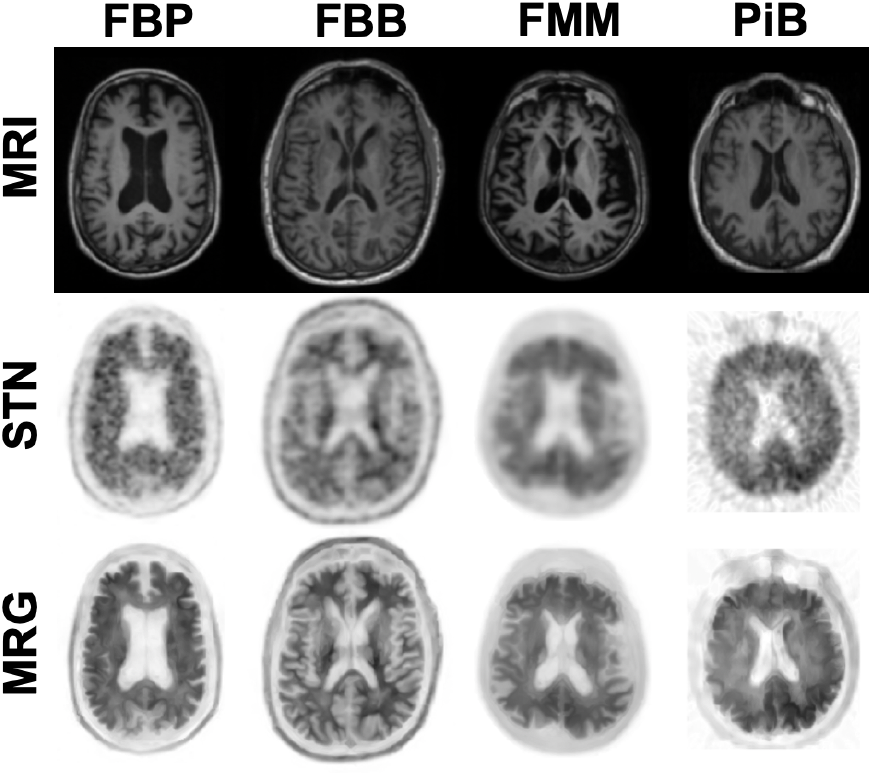
STN PET and MRG PET across tracers. Structural T1-weighted MRI is used to guide enhancement of the standard PET reconstruction (STN), producing the MRI-guided PET output (MRG) for Florbetapir (FBP), Florbetaben (FBB), Flutemetamol (FMM), and Pittsburgh Compound B (PiB).

**Fig. 2:**
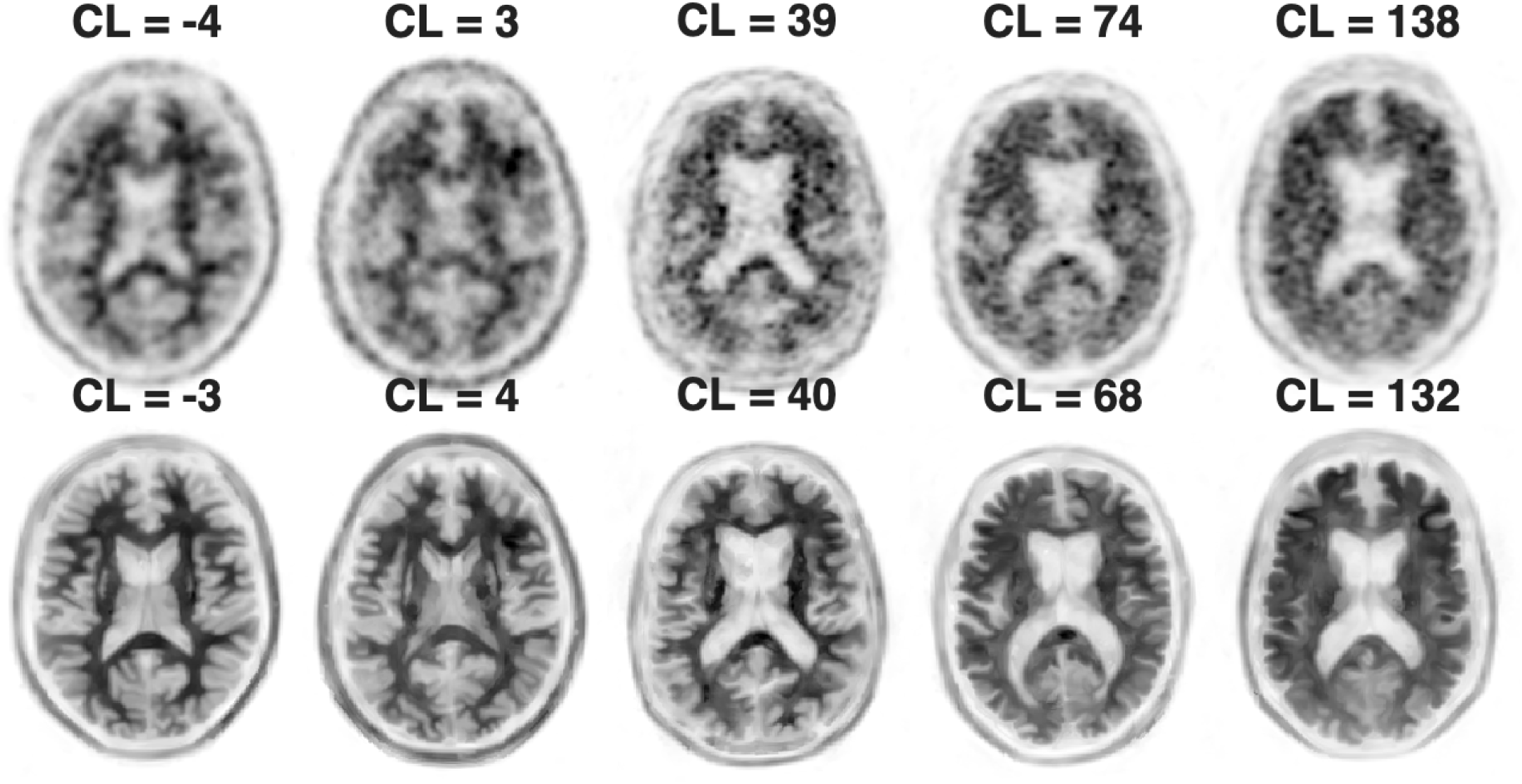
STN and MRG Florbetapir PET across independently calibrated Centiloid scores. Standard PET images are shown in the top row and corresponding MR-guided PET denoising and resolution enhancement outputs are shown in the bottom row. Centiloid values were calibrated separately for STN and MRG using processing-condition-specific mappings to the GAAIN Centiloid reference framework. MRG improves cortical definition across a broad range of amyloid burden while preserving the overall tracer uptake pattern.

**Fig. 3:**
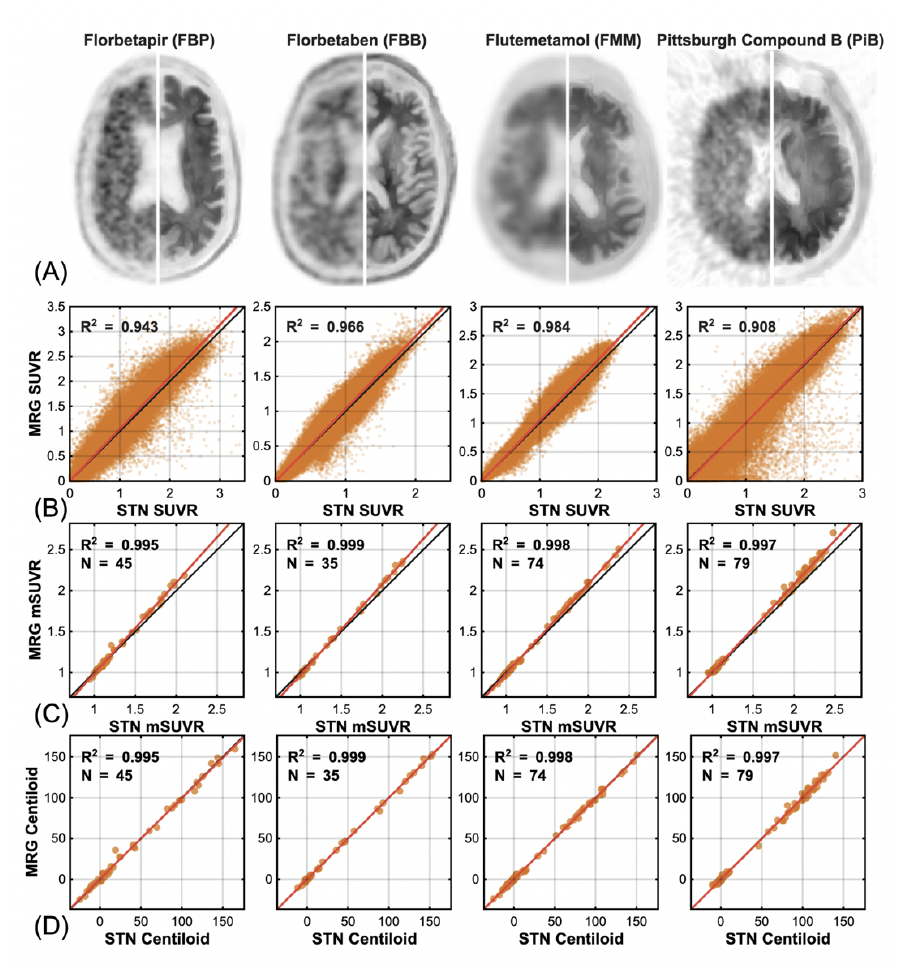
SUVr agreement between STN and MRG. A) Top: Representative individual Voxelwise SUVr correlations show high correlations between STN SUVr and MRG SUVr. Example images displaying STN PET on the left and MRG PET on the right for FBB, FBP, FMM and PiB. Bottom: Voxelwise correlation plots between STN SUVr and MRG SUVr for FBP (*R*^2^ = 0.943), FBB (*R*^2^ = 0.966), FMM (*R*^2^ = 0.984), and PiB (*R*^2^ = 0.908). B) Group Median CTX SUVr correlations show high correlations between STN SUVr and MRG SUVr. SUVr correlation plots between STN SUVr and MRG SUVr for FBP (*R*^2^ = 0.995), FBB (*R*^2^ = 0.999), FMM (*R*^2^ = 0.998), and PiB (*R*^2^ = 0.997). C) Centiloid correlations show high correlations between STN Centiloid and MRG Centiloid. Centiloid correlation plots between STN Centiloid and MRG Centiloid for FBP (*R*^2^ = 0.995), FBB (*R*^2^ = 0.999), FMM (*R*^2^ = 0.998), and PiB (*R*^2^ = 0.997).

**Fig. 4:**
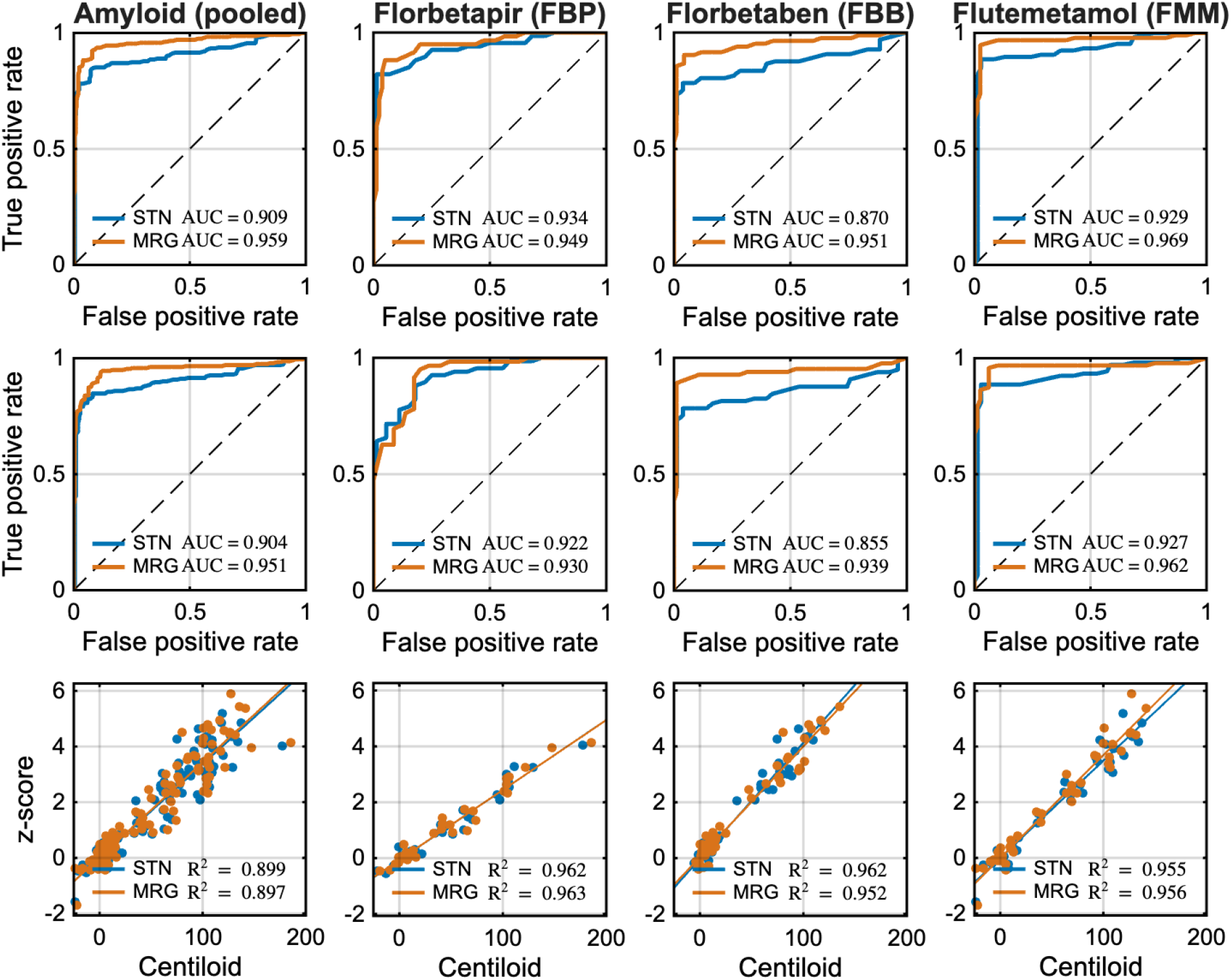
Concordance Analysis between raters and Centiloid and z-scores. ROC curves show concordance for STN (orange) and MRG (blue) between binary amyloid tracer positivity reads and quantitative metrics. A) ROC curves for Centiloid values for pooled amyloid cases (AUC: STN = 0.909, MRG = 0.959), and FBP (AUC: STN = 0.934, MRG = 0.949), FBB (AUC: STN = 0.870, MRG = 0.951), and FMM (AUC: STN = 0.929, MRG = 0.969), comparing STN and MRG. B) ROC curves for z-score values for pooled amyloid cases (AUC: STN = 0.904, MRG = 0.951), and FBP (AUC: STN = 0.922, MRG = 0.930), FBB (AUC: STN = 0.855, MRG = 0.939), and FMM (AUC: STN = 0.927, MRG = 0.962), comparing STN and MRG. C) Correlation plots between Centiloid and z-score values for pooled amyloid tracers (STN: *R*^2^ = 0.899, MRG: *R*^2^ = 0.897), FBP (STN: *R*^2^ = 0.962, MRG: *R*^2^ = 0.963), FBB (STN: *R*^2^ = 0.962, MRG: *R*^2^ = 0.952) and FMM (STN: *R*^2^ = 0.955, MRG: *R*^2^ = 0.956) for STN and MRG.

Data used in the preparation of this article were obtained in part from the Alzheimer’s Disease Neuroimaging Initiative (ADNI) database (https://adni.loni.usc.edu). ADNI was launched in 2003 as a public-private partnership, led by Principal Investigator Michael W. Weiner, MD. The original goal of ADNI was to test whether serial magnetic resonance imaging, positron emission tomography, other biological markers, and clinical and neuropsychological assessment could be combined to measure the progression of mild cognitive impairment and early Alzheimer’s disease. Current ADNI goals include validating biomarkers for clinical trials, improving the generalizability of ADNI data by increasing diversity in the participant cohort, and providing data concerning the diagnosis and progression of Alzheimer’s disease to the scientific community. ADNI data used in this study were downloaded from the LONI Image and Data Archive across multiple sessions between July 2025 and January 2026, and the database was checked for updated data prior to manuscript submission.

Quantitative analyses used subjects from the GAAIN Centiloid Project reference data. These datasets included both MR and PET images for each subject and contained subject imaging from four different amyloid PET tracers, including [^11^C]Pittsburgh compound-B (PiB, n = 79), [^18^F]AV-45 ([^18^F]florbetapir, FBP, n = 45), [^18^F]florbetaben (FBB, n = 35), and [^18^F]flutemetamol (FMM, n = 74). Each subject from FBP, FBB and FMM datasets had an additional PiB image to satisfy calibration requirements. PiB data were used for quantitative analyses only and were not included in the blinded reader study. Patient demographic information is described in detail in Table 1. Images used in the quantitative analyses were taken from eight PET scanners and 12 MRI scanners (Supp. Table 1).

**Table 1:** Demographics and amyloid tracer types of GAAIN datasets for each of the four amyloid tracers for different diseased and healthy control cohorts: Alzheimer’s disease (AD), mild cognitive impairment (MCI), frontotemporal dementia (FTD), Elderly (E) and young control (YC).

| Tracer | PiB | FBP | FBB | FMM |
| --- | --- | --- | --- | --- |
| **AD (n)** | 45 | 17 | 8 | 20 |
| Age | 68 ± 11 | 67 ± 7 | 69 ± 6 | 69 ± 7 |
| M/F | N/A | 8/9 | 4/4 | 6/14 |
| **MCI (n)** | - | 6 | 9 | 20 |
| Age | - | 79 ± 9 | 72 ± 5 | 73 ± 7 |
| M/F | - | 5/1 | 4/5 | 11/9 |
| **FTD (n)** | - | - | 2 | - |
| Age | - | - | 74 ± 8 | - |
| M/F | - | - | 2/1 | - |
| **E (n)** | - | 9 | 6 | 10 |
| Age | - | 69 ± 11 | 72 ± 9 | 57 ± 11 |
| M/F | - | 7/2 | 2/4 | 3/7 |
| **YC (n)** | 34 | 13 | 10 | 24 |
| Age | 32 ± 6 | 27 ± 4 | 33 ± 9 | 37 ± 5 |
| M/F | N/A | 6/7 | 3/7 | 10/14 |
| **Total n** | 79 | 45 | 35 | 74 |

The blinded multi-reader study used subjects assembled from GAAIN, the Alzheimer’s Disease Neuroimaging Initiative (ADNI), and GE HealthCare. Reader analyses were performed in tracer-specific FBP (n=35), FBB (n=44), and FMM (n=44) cohorts with age and sex-matched subjects. Patient demographic and data source information is described in Table 2. Images used in the blinded reader study were taken from 33 PET scanners and 22 MRI scanners (Supp. Table 2).

**Table 2:** Demographics for GAAIN, GE and ADNI datasets used in the rater study include diseased and healthy control cohorts: Alzheimer’s disease (AD), mild cognitive impairment (MCI), and cognitively normal (CN).

| Tracer | FBP | FBB | FMM |
| --- | --- | --- | --- |
| **AD/MCI** | 13 | 22 | 34 |
| Age | 78 ± 5 | 76 ± 7 | 75 ± 4 |
| M/F | 8/5 | 11/11 | 17/17 |
| **CN** | 22 | 22 | 10 |
| Age | 75 ± 6 | 74 ± 5 | 58 ± 11 |
| M/F | 11/11 | 11/11 | 3/7 |
| Data source | ADNI | ADNI | GE/GAAIN |

### 2.2 Ethics approval and consent

This study used de-identified retrospective imaging data from ADNI, GAAIN, and GE HealthCare datasets in accordance with applicable data-use agreements. No prospective participant recruitment or intervention was performed, and no new informed consent was required for this secondary analysis.

### 2.3 STN-PET reconstruction and MRG-PET denoising and resolution enhancement

Amyloid PET acquisition protocols were harmonized across tracers using Centiloid framework standards and reference datasets from the GAAIN Centiloid Project for STN-PET reconstruction. For the GAAIN Centiloid Project reference data, PiB scans followed the standard 50-70 min post-injection acquisition window defined by the Centiloid Level 1–2 reference datasets [25]. GAAIN FBP, FBB, and FMM datasets were processed using tracer-specific acquisition windows aligned to the Centiloid framework, with 50-60 min for FBP, and 90-110 min for FBB and FMM [33– 36]. For the reader-study data, ADNI FBP images were generated by averaging four 5-minute frames acquired over 50-70 min after motion correction and co-registration, ADNI FBB used a 20-minute static acquisition beginning at approximately 90 min post-injection, and GE HealthCare FMM used a 30 minute acquisition ∼ 90 min after injection with static images generated from motion-corrected dynamic frames [31, 32]. Structural T1-weighted MRI was available for all subjects for subsequent MRG processing. MRI and PET were not required to have been acquired on the same scanner or during the same imaging session, and MRG is MR contrast-agent agnostic. Standard clinical PET reconstructions from each source dataset were treated as the standard-of-care (STN) images used as input for subsequent comparative analyses.

### 2.4 Image processing and MR-guided denoising and resolution enhancement

STN PET images were enhanced with structural MRI using MR-guided denoising and resolution enhancement (MRG), (Figs. 1, 2). Each STN PET image was first affinely co-registered to the corresponding structural T1-weighted MRI in native space using mutual information as the similarity metric. MRG PET images were obtained by solving a convex optimization problem consisting of a PET data-fidelity term and an MRI-guided Bowsher regularization term. The data-fidelity term constrained the solution to remain consistent with the input PET image, while the Bowsher term used local anatomical similarity in the MRI to regularize the PET image according to underlying structural boundaries. Optimization was performed using a Nesterov-accelerated iterative scheme, and the regularization weight, *β*, was estimated automatically based on PET noise level and point-spread function (Supp. Fig. 1). MRG was applied as a post-reconstruction image-processing step to reconstructed PET DICOM images and did not require access to raw PET sinogram data, scanner-specific reconstruction software, or segmentation-driven partial volume correction.

### 2.5 Amyloid regions of interest & SUVr

Quantitative analyses used amyloid regions of interest (ROIs) defined by the GAAIN Centiloid Project. ROI definitions and reference-region selection followed GAAIN Centiloid Project conventions for amyloid PET quantification. ROIs provided in MNI-152 space were registered into each subject’s native space before quantitative measurement using proprietary software. The Whole Cerebellum was used as the reference ROI, consistent with the standard PiB Centiloid framework [25]. The cortical target region (CTX) included the precuneus, anterior striatum, and frontal, temporal, parietal, and insular cortices. Standardized Uptake Value Ratio (SUVr) was computed for both STN and MRG images as the ratio of median uptake within the CTX to median uptake within the Whole Cerebellum reference ROI. Median uptake was used rather than mean uptake to reduce sensitivity to outlier voxel values.

### 2.6 Centiloid Scaling

Centiloid values were derived using the GAAIN Centiloid reference framework. SUVr values produced by the present processing pipeline were not expected to numerically match those generated by the GAAIN reference implementation, even when representing the same underlying tracer burden, due to differences in image processing and quantification. Direct calibration was therefore performed before Centiloid conversion. For each tracer, including PiB, FBP, FBB, and FMM, tracer-specific linear mappings were fit from measured SUVr values to the corresponding GAAIN reference values. These mappings were performed separately for STN and MRG images and were then used to express each processing condition on the GAAIN-aligned Centiloid scale. This separate calibration was performed because MRG alters the effective spatial resolution of the PET image, and recent work has shown that applying the same Centiloid conversion equations across images with different spatial resolution can introduce systematic bias [37]. For PiB, this step aligned our processed PiB SUVr values to the GAAIN PiB reference implementation, while for FBP, FBB, and FMM it provided tracer-specific Level 2 calibration to the GAAIN reference framework.

### 2.7 SUVr z-Score

SUVr-derived z-scores were computed using tracer-specific tracer-negative reference cohorts from the GAAIN young control datasets. Reference cohort sizes were n = 34 for PiB, n = 13 for FBP, n = 10 for FBB, and n = 24 for FMM. For each tracer, subject SUVr images were compared against the corresponding tracer-negative reference distribution, and the median z-score within the CTX was used as the summary measure. In contrast to Centiloid scaling, which maps tracer uptake onto a standardized anchored scale, z-score quantification reflects the number of standard deviations from the tracer-negative reference mean and is therefore unbounded. Z-score was included as a complementary quantitative measure of abnormal tracer burden.

### 2.8 Blinded multi-reader study

Four board-certified radiologists with 8+ years of experience interpreting brain PET independently evaluated anonymized STN and MRG images in a blinded multi-reader design. Readers were informed of tracer type but were blinded to image origin and to all quantitative data. STN and MRG images were presented on separate standardized slides, with one processing condition per slide and five axial and five coronal slices shown for each case. Image review was performed using standardized slide decks distributed as PDFs rather than within a clinical PACS environment, in order to ensure consistent presentation across readers and maintain blinding to processing condition. Reads were performed separately by tracer to preserve tracer-appropriate interpretation. Qualitative endpoints included image quality, artifact severity, reader confidence, and binary amyloid tracer positivity (positive/negative). Image quality was rated on a 5-point ordinal scale with “5” being excellent, artifact severity and reader confidence were rated on a 3-point ordinal scale with “3” being no artifact, and “3” being highly confident. Amyloid burden was additionally categorized by the reader as sparse, mild, moderate or severe for burden-stratified analyses. Examples of varying image quality, artifacts, and disease severity were included in reader instructions to standardize ratings across readers.

### 2.9 Quantitative and reader-performance statistical analyses

Quantitative preservation of MRG relative to STN was evaluated by comparing matched SUVr and Centiloid measurements across amyloid tracers. Representative voxel-wise SUVr comparisons were used to assess preservation of the spatial uptake distribution, and cohort-level comparisons used median CTX SUVr and derived Centiloid values from each subject. Quantitative agreement between STN and MRG was summarized using linear regression and coefficients of determination (*R*^2^) (Fig. 3). To assess visual-quantitative concordance, receiver operating characteristic (ROC) curves were generated using qualitative reader amyloid-positivity labels as the binary reference and continuous Centiloid or z-score values as the test variable. Area under the curve (AUC) was summarized separately for STN and MRG images for pooled amyloid and for each tracer (Fig. 4).

Tracer-specific correlations between Centiloid and z-score were evaluated as an internal consistency check between complementary quantitative scales. Reader-performance analyses were used to assess how MRG affected reader behavior. Inter-rater agreement was quantified as the mean pairwise agreement across readers within each STN and MRG image, defined as the proportion of agreeing reader pairs among all possible reader pairs. Fleiss’ kappa was computed to assess chance-corrected multi-reader reliability. Agreement and Fleiss’ kappa were summarized with 95% confidence intervals estimated by bootstrap resampling at the subject level; bootstrap kappa estimates were constrained to valid values (-1 to 1), and estimates with near-zero or non-finite denominators were excluded to prevent numerical instability.

Because external ground-truth labels were not available, reader performance accuracy was also evaluated using a leave-one-reader-out bronze standard – for each read, the reference label was defined as the majority vote of the other readers evaluating the same subject under the same processing condition. Each reader’s classification was then compared against this bronze-standard label to determine correctness. Bronze-standard performance was summarized as accuracy, false-positive (FP) rate, and false-negative (FN) rate. Rates were computed by aggregating counts across reads within each tracer and processing condition, with FP rate defined as FP / (TN + FP) and FN rate defined as FN / (TP + FN). Confidence intervals for agreement, accuracy, FP rate, and FN rate were estimated using subject-level bootstrap resampling to account for within-case clustering (Fig. 5).

**Fig. 5:**
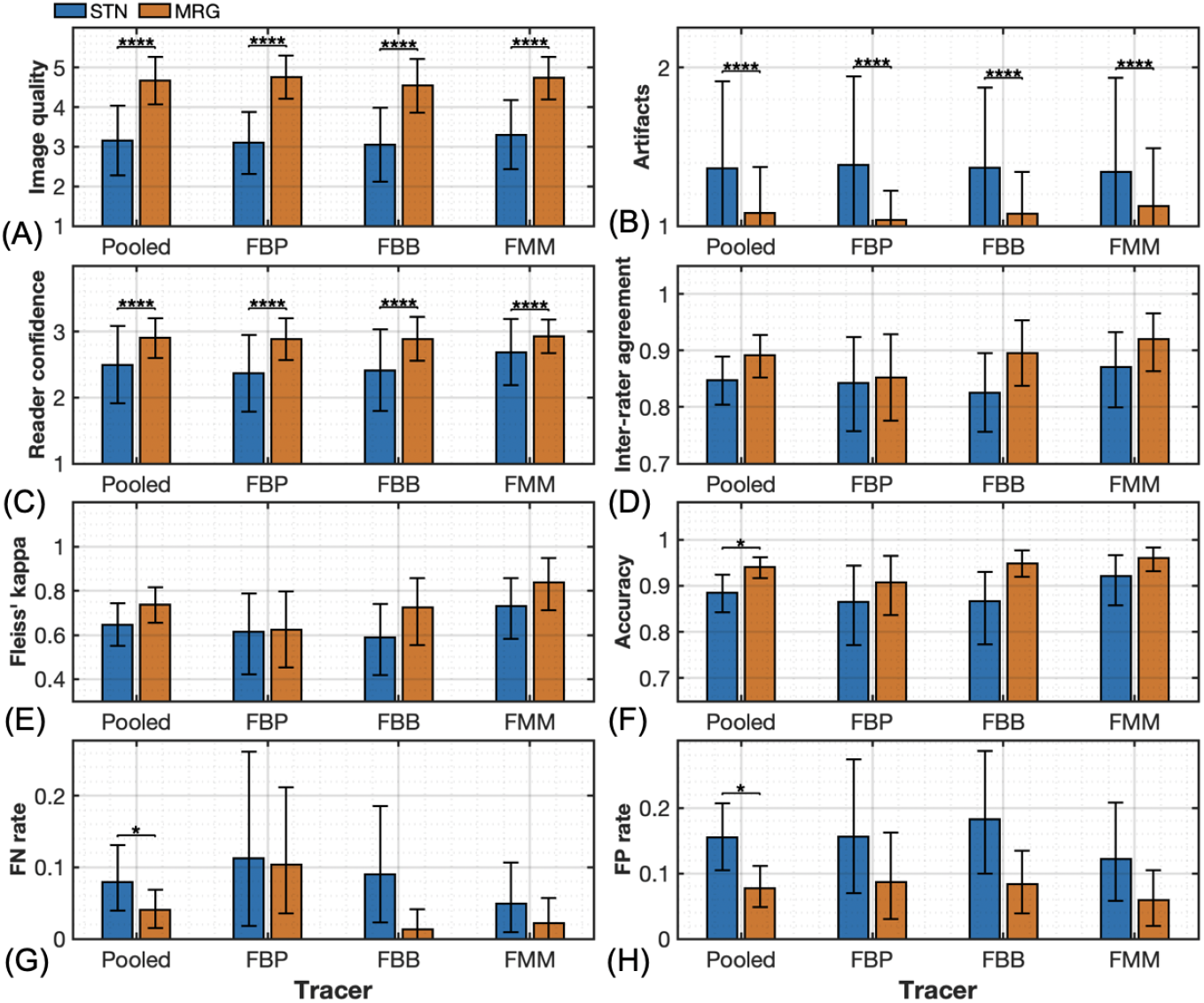
MRG increases reader interpretability and reliability of amyloid PET images across tracers in the blinded reader study. Comparisons of STN (orange) and MRG (blue) in qualitative reader endpoints and reader performance metrics from the blinded reader study separated by tracer. A) Image quality ratings for pooled amyloid, and individual tracers FBP, FBB and FMM (all *p* < 0.0001, paired Wilcoxon Signed-Rank). B) Artifact ratings for pooled amyloid, FBP, FBB and FMM (all *p* < 0.0001, paired Wilcoxon Signed-Rank). C) Reader confidence comparisons for pooled amyloid, FBP, FBB, and FMM (all *p* < 0.0001, paired Wilcoxon Signed-Rank). D) Interrater agreement comparisons for pooled amyloid (*p* = 0.080), FBP (*p* = 0.555), FBB (*p* = 0.165), and FMM (*p* = 0.302, all paired Wilcoxon Signed-Rank). E) Fleiss’ kappa for rater-agreement using pooled amyloid, FBP, FBB, and FMM. F) Accuracy for pooled amyloid, FBP, FBB, and FMM. G) False Negative Rate for pooled amyloid, FBP, FBB, and FMM. H) False Positive Rate for pooled amyloid, FBP, FBB, and FMM.

Additional analyses examined whether the effect of MRG varied across amyloid burden. Cases were stratified using predefined Centiloid and z-score bins, and reader-derived metrics, including confidence, agreement, accuracy, and FN rate, were summarized within each burden range (Fig. 6). Quantitative values used for these stratified analyses were taken from the STN reconstruction for both STN and MRG reads to preserve a common reference scale across processing conditions. Centiloid bins were defined as follows: low burden: < 25, moderate burden 25-100, and high burden 100+. Z-score bins were defined as follows: low burden: < 1, moderate burden: 1-3, high burden 3+. Within each bin, summary statistics were computed separately for STN and MRG and reported with 95% confidence intervals estimated by subject-level bootstrap resampling. Ordinal reader endpoints, (image quality, artifact severity, and reader confidence) and reader performance metrics (inter-rater agreement, accuracy, FP rate, and FN rate) were compared between STN and MRG using paired Wilcoxon Signed-Rank tests (Fig. 5). Accuracy stratified by Centiloid and z-score were compared using paired Wilcoxon Signed-Rank. Reader Confidence, Accuracy, and Centiloid score stratified by reader severity score, and FN rate stratified by Centiloid and z-score were compared using Wilcoxon Signed-Rank tests, where paired STN/MRG observations were not available (Fig. 6).

**Fig. 6:**
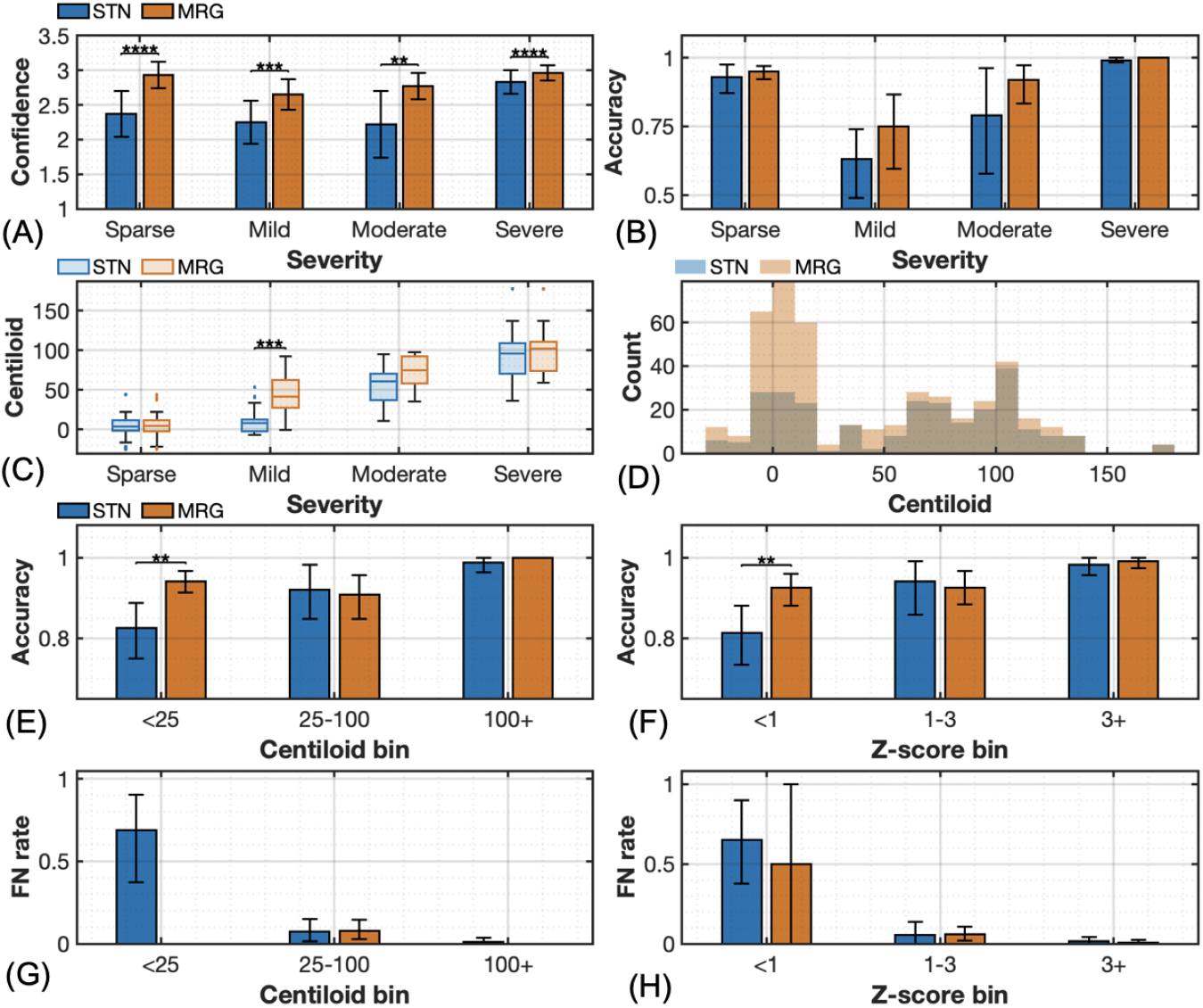
MRG improves reader ability to distinguish early-stage amyloid burden as compared to STN. Comparisons of STN (orange) and MRG (blue) in qualitative reader endpoints and reader performance metrics from the blinded reader study stratified by disease severity, Centiloid or z-score. A) Reader confidence ratings stratified by disease severity; sparse (*p* < 0.0001), mild (*p* < 0.001), moderate (*p* < 0.01), and severe (*p* < 0.0001, all Wilcoxon Signed-Rank). B) Accuracy stratified by disease severity; sparse (*p* = 0.848), mild (*p* = 0.113), moderate (*p* = 0.666), and severe (*p* = 0.363, all Wilcoxon Signed-Rank). C) Centiloid scores stratified by disease severity sparse (*p* = 0.865), mild (*p* < 0.001), moderate (*p* = 0.101), and severe (*p* = 0.615, all Wilcoxon Signed-Rank). D) Histogram of confidence = 3 scores distributed by Centiloid score for STN and MRG. E) Accuracy stratified by Centiloid score; < 25 (*p* < 0.01), 25–100 (*p* = 0.59), 100+ (*p* = 1, all paired Wilcoxon Signed-Rank). F) Accuracy stratified by z-score; < 1 (*p* < 0.01), 1–3 (*p* = 0.562), 3+ (*p* = 1, all paired Wilcoxon Signed-Rank). G) False Negative Rate stratified by Centiloid score; < 25 (*p* = NaN), 25–100 (*p* = 0.36), 100+ (*p* = 0.34, all Wilcoxon Signed-Rank). H) False Negative Rate stratified by z-score; < 1 (*p* = 0.939), 1–3 (*p* = 0.267), 3+ (*p* = 0.57, all Wilcoxon Signed-Rank).

## 3 Results

### 3.1 MRG-PET improves cortical conspicuity across tracers and amyloid burden levels

MR-guided denoising and resolution enhancement (MRG) improved cortical conspicuity relative to standard reconstruction (STN) across all four amyloid tracers (Fig. 1). In representative cases, MRG increased visible separation between cortical gray matter and adjacent white matter interfaces, while preserving the overall tracer uptake pattern relative to STN, (Figs. 1–2). Additional FBP examples spanning low (3) to high (185) Centiloid Scores show that this apparent spatial sharpening was present over a broad range of amyloid burden (Fig. 2).

### 3.2 MRG preserves quantitative amyloid PET metrics relative to STN

MRG preserved established quantitative amyloid PET metrics, measured through SUVr and Centiloid, after calibration to STN. Voxel-wise SUVr correlation between STN and MRG in representative subjects on each amyloid tracer cohort show a tight clustering around the identity line. This is supported by high *R*^2^ values, FBP (0.943), FBB (0.966), FMM (0.984), PiB (0.908), (Fig. 3A). At the population level, median CTX SUVr values from each subject remained strongly correlated between STN and MRG across tracers, FBP (0.995), FBB (0.999), FMM (0.998), PiB (0.997), (Fig. 3B). These same *R*^2^ values were observed for Centiloid values after tracer-specific calibration (Fig. 3C), supporting quantitative agreement of the MRG pipeline to STN for both region-based SUVr and derived Centiloid measures. MRG did not introduce bias into group SUVr metrics, as demonstrated by low mean differences in SUVr measurements (MRG – STN: FBP=0.025, FBB=0.016, FMM=0.019, PiB=0.051). This corresponded to only modest percent increases in MRG relative to STN (FBP=1.9%, FBB=1.1%, FMM=1.4%, PiB=3.2%).

The modest systematic increase in SUVr observed with MRG likely reflects reduced partial volume effects due to improved effective resolution. As the point spread function broadens, signal is mixed between gray and white matter, reducing cortical uptake values with dilution of the surrounding signal; reducing this blur shifts these values in the opposite direction, which can increase ROI-based SUVr depending on boundary definition (Supplemental Fig. 1). Importantly, these shifts do not affect Centiloid values, which are calibrated to a common reference dataset (GAAIN).

### 3.3 MRG improves concordance between visual interpretation and quantitative amyloid burden

Because MRG preserved quantitative metrics, we next tested whether it improved concordance between visual reads and quantitative measures of amyloid burden across FBP, FBB, and FMM using reader-defined tracer positivity labels and continuous Centiloid or z-score values. ROC analyses showed higher AUC for MRG than for STN for pooled amyloid and for each individual tracer for both Centiloid (MRG: pooled: 0.959, FBP: 0.949, FBB: 0.951, FMM: 0.969; STN: pooled: 0.909, FBP: 0.934, FBB: 0.870, FMM: 0.929) and z-score (MRG: pooled: 0.951, FBP: 0.930, FBB: 0.939, FMM: 0.962; STN: pooled: 0.904, FBP: 0.922, FBB: 0.855, FMM: 0.927), (Fig. 4A, B). These findings indicate higher concordance between visual amyloid positivity and quantitative uptake metrics with MRG processing as compared to STN. A high correlation between Centiloid and z-score was preserved for both MRG and STN methods across FBP, FBB and FMM (*R*^2^: MRG: pooled: 0.897, FBP: 0.963, FBB: 0.952, FMM: 0.956; STN: pooled: 0.899, FBP: 0.962, FBB: 0.962, FMM: 0.955), supporting that both quantitative measures are internally consistent and track the same underlying burden scale (Fig. 4C) in both STN and MRG.

### 3.4 MRG improves qualitative image ratings, image interpretability, and reader performance across amyloid PET tracers

MRG improved qualitative reader endpoints and multi-reader performance in the blinded reader study (Fig. 5). Across amyloid tracers, MRG was associated with significantly higher image quality ratings, lower artifact burden, and higher reader confidence than STN in pooled amyloid and in each individual tracer cohort (Fig. 5A–C; all *p* < 0.0001). Pooled average image quality was 4.7 for MRG and 3.2 for STN (ordinal scale 1-5, with “5” being excellent); pooled average artifact burden values out of 3 were 1.1 for MRG and 1.4 for STN (ordinal scale 1-3 with “3” being no artifact); pooled average reader confidence values out of 3 were 2.9 for MRG and 2.5 for STN (ordinal scale 1-3 with “3” being highly confident). Individual tracers followed these trends for all three reader performance metrics (Table 3), displaying improvement across all metrics with MRG as compared to STN. These findings indicate that MRG enhanced the visual interpretability of amyloid PET images across tracers.

**Table 3:** Qualitative reader results across amyloid tracers. Results for qualitative readers endpoints (Fig. 5A–C) listed as mean*±*sd for pooled amyloid and across individual amyloid tracers for STN and MRG processing.

| Tracer | Pooled | FBP | FBB | FMM |
| --- | --- | --- | --- | --- |
| STN Image Quality | 3.20 ± 0.88 | 3.10 ± 0.78 | 3.05 ± 0.94 | 3.30 ± 0.87 |
| MRG Image Quality | 4.70 ± 0.60 | 4.76 ± 0.55 | 4.54 ± 0.68 | 4.73 ± 0.54 |
| STN Artifact | 1.40 ± 0.55 | 1.39 ± 0.56 | 1.37 ± 0.51 | 1.34 ± 0.59 |
| MRG Artifact | 1.10 ± 0.29 | 1.04 ± 0.19 | 1.08 ± 0.27 | 1.12 ± 0.36 |
| STN Confidence | 2.50 ± 0.58 | 2.37 ± 0.58 | 2.41 ± 0.62 | 2.69 ± 0.50 |
| MRG Confidence | 2.90 ± 0.30 | 2.89 ± 0.32 | 2.89 ± 0.33 | 2.93 ± 0.25 |

Inter-rater agreement did not differ significantly between STN and MRG by mean pairwise agreement (Fig. 5D; pooled *p* = 0.080, FBP *p* = 0.555, FBB *p* = 0.165, FMM *p* = 0.302),although pooled average values for STN were 0.85 and improved to 0.89 for MRG and this same trend was observed across all individual tracers (Table 4). Fleiss’ kappa values were generally higher with MRG than with STN, including in pooled amyloid (MRG *κ* = 0.74 [0.65–0.82] vs STN *κ* = 0.65 [0.55–0.74]), with the same overall directional pattern observed across FBP, FBB, and FMM (Fig. 5E, Table 4). This suggests improved chance-corrected multi-reader reliability of amyloid positivity assessments with MRG.

**Table 4:** Reader performance results across amyloid tracers. Results for reader performance metrics (Fig. 5D–G) listed as mean [95% CI] for pooled amyloid and across individual amyloid tracers for STN and MRG processing.

| Tracer | Pooled | FBP | FBB | FMM |
| --- | --- | --- | --- | --- |
| STN Inter-rater Agreement | 0.85 [0.80, 0.89] | 0.84 [0.76, 0.92] | 0.83 [0.76, 0.90] | 0.87 [0.80, 0.93] |
| MRG Inter-rater Agreement | 0.89 [0.85, 0.93] | 0.85 [0.78, 0.93] | 0.90 [0.84, 0.95] | 0.92 [0.86, 0.97] |
| STN Fleiss Kappa | 0.65 [0.55, 0.74] | 0.61 [0.42, 0.79] | 0.59 [0.42, 0.74] | 0.73 [0.58, 0.86] |
| MRG Fleiss Kappa | 0.74 [0.65, 0.82] | 0.62 [0.45, 0.80] | 0.72 [0.55, 0.86] | 0.84 [0.71, 0.95] |
| STN Accuracy | 0.89 [0.84, 0.92] | 0.86 [0.77, 0.94] | 0.87 [0.77, 0.93] | 0.92 [0.86, 0.97] |
| MRG Accuracy | 0.94 [0.92, 0.96] | 0.91 [0.84, 0.96] | 0.95 [0.92, 0.98] | 0.96 [0.93, 0.98] |
| STN FN Rate | 0.08 [0.04, 0.13] | 0.11 [0.02, 0.26] | 0.09 [0.02, 0.19] | 0.05 [0.01, 0.11] |
| MRG FN Rate | 0.04 [0.02, 0.07] | 0.10 [0.04, 0.21] | 0.01 [0.00, 0.04] | 0.02 [0.00, 0.06] |
| STN FP Rate | 0.15 [0.10, 0.21] | 0.16 [0.07, 0.27] | 0.18 [0.10, 0.29] | 0.12 [0.06, 0.21] |
| MRG FP Rate | 0.08 [0.05, 0.11] | 0.09 [0.03, 0.16] | 0.08 [0.04, 0.13] | 0.06 [0.02, 0.11] |

Improvements in image interpretability were accompanied by favorable shifts in reader performance metrics. Relative to STN, MRG showed greater accuracy (Fig. 5F, pooled: *p* = 0.018, FBP: *p* = 0.555, FBB: *p* = 0.057, FMM: *p* = 0.241) with average pooled values increasing from 0.89 to 0.94, and similar shifts in trends for individual tracers. MRG showed an average pooled False Negative Rate of 0.04, while it was 0.08 for STN, a pattern that was followed by individual tracers (Table 4) (Fig. 5G, pooled: *p* = 0.047, FBP: *p* = 0.625, FBB: *p* = 0.125, FMM: *p* = 0.531). Similarly, MRG showed an average pooled False Positive Rate of 0.08, which increased to 0.15 for STN with similar trends across all individual tracers (Table 4) (Fig. 5H, pooled: *p* = 0.016, FBP: *p* = 0.250, FBB: *p* = 0.127, FMM: *p* = 0.388). These effects reached significance in pooled amyloid analyses, with similar directional trends across individual tracers. Together, these findings indicate that the observed visual improvements with MRG were associated with more consistent and higher-accuracy binary reader interpretation.

### 3.5 MRG has the strongest effect on reader confidence and accuracy in visually challenging low-to-intermediate amyloid-burden PET cases

To determine whether the effect of MRG varied across amyloid burden, cases were first stratified by reader severity classification, independent of tracer type. Reader confidence was higher with MRG than with STN for sparse (*p* < 0.0001, STN: *n* = 41, MRG: *n* = 61), mild (*p* < 0.001, STN: *n* = 23, MRG: *n* = 13), moderate (*p* < 0.01, STN: *n* = 13, MRG: *n* = 10), and severe cases (*p* < 0.0001, STN: *n* = 46, MRG: *n* = 40), improving from 2.4 to 2.9, 2.3 to 2.7, 2.2 to 2.8, and 2.8 to 3.0, respectively (Fig. 6A, Table 5). There were no significant differences between MRG and STN when accuracy was stratified by reader severity classification, although both mild and moderate ratings trended towards improved accuracy with MRG (Fig. 6B). Mild increased from 0.63 to 0.75 (*p* = 0.113, STN: *n* = 22, MRG: *n* = 13), and moderate increased from 0.79 to 0.92 (*p* = 0.666, STN: *n* = 13, MRG: *n* = 9), while sparse increased from 0.93 to 0.95 (*p* = 0.848, STN: *n* = 41, MRG: *n* = 60) and severe from 0.99 to 1.0 (*p* = 0.363, STN: *n* = 46, MRG: *n* = 40) (Table 5). These findings indicate that the reader-facing benefit of MRG was most apparent in lower amyloid burden cases that are the most visually challenging to interpret in routine clinical care.

**Table 5:**
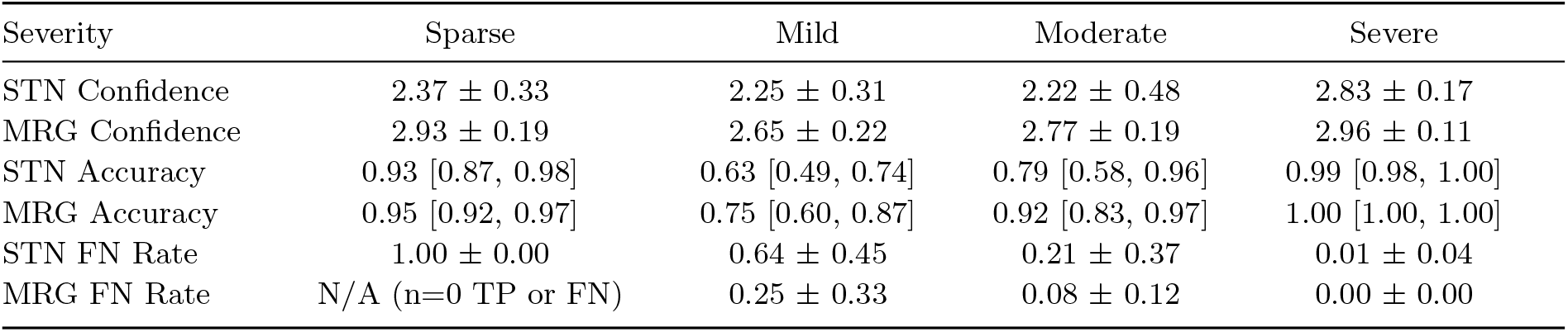
Confidence and Accuracy for STN and MRG stratified by reader designated disease severity ratings. Results for Reader Confidence (Fig. 6A) listed as mean*±*sd, Accuracy (Fig. 6B) listed as mean [95% CI], and False Negative Rate (not included in Fig. 6) for sparse, mild, moderate, and severe disease burden for STN and MRG processing.

| Severity | Sparse | Mild | Moderate | Severe |
| --- | --- | --- | --- | --- |
| STN Confidence | 2.37 ± 0.33 | 2.25 ± 0.31 | 2.22 ± 0.48 | 2.83 ± 0.17 |
| MRG Confidence | 2.93 ± 0.19 | 2.65 ± 0.22 | 2.77 ± 0.19 | 2.96 ± 0.11 |
| STN Accuracy | 0.93 [0.87, 0.98] | 0.63 [0.49, 0.74] | 0.79 [0.58, 0.96] | 0.99 [0.98, 1.00] |
| MRG Accuracy | 0.95 [0.92, 0.97] | 0.75 [0.60, 0.87] | 0.92 [0.83, 0.97] | 1.00 [1.00, 1.00] |
| STN FN Rate | 1.00 ± 0.00 | 0.64 ± 0.45 | 0.21 ± 0.37 | 0.01 ± 0.04 |
| MRG FN Rate | N/A (n=0 TP or FN) | 0.25 ± 0.33 | 0.08 ± 0.12 | 0.00 ± 0.00 |

A similar effect was observed when cases were stratified by quantitative amyloid burden. Reader severity classification tracked closely with Centiloid values (Fig. 6C; sparse: *p* = 0.866, moderate: *p* = 0.101, severe: *p* = 0.615), while mild cases had higher Centiloid values with MRG than STN (*p* < 0.001). High-confidence reads were more frequent with MRG (*n* = 441) than with STN (*n* = 265), with most of this difference occurring in the lower Centiloid range (− 10 to 30) (Fig. 6D). Accuracy was 0.83 with STN and increased to 0.94 with MRG for lowest Centiloid bin (Fig. 6E, *p* < 0.01, *n* = 60) and was 0.81 with STN and increased to 0.92 with MRG for the lowest z-score bin (Fig. 6F, *p* < 0.01, *n* = 63), with no statistical differences within the two higher burden bins (Centiloid: 25–100: *p* = 0.590, *n* = 41, 100+: *p* = 1, *n* = 21; z-score: 1–3: 0.563, *n* = 30, 3+: *p* = 1, *n* = 29). False-negative reads in the low-burden bin were reduced from 0.69 with STN to 0 with MRG in the lowest Centiloid bin (Fig. 6G, MRG: *n* = 0, STN: *n* = 8) and a partial reduction in the lowest z-score bin from 0.65 with STN to 0.50 with MRG (Fig. 6H, MRG: *n* = 2, STN: *n* = 10), with lesser reductions with MRG in the two higher bins for both Centiloid and z-score (Tables 6, 7). These findings emphasize that a major benefit of MRG is improved confidence and accuracy for low-burden amyloid PET cases near the visual positivity threshold.

**Table 6:** Accuracy and False Negative Rate for STN and MRG stratified by Centiloid score. Reader Confidence (not included in Fig. 6) listed as mean*±*sd, Accuracy (Fig. 6E) and False Negative Rate (Fig. 6G) listed as mean [95% CI] across three Centiloid bins for STN and MRG processing.

| Centiloid | < 25 | 25–100 | 100+ |
| --- | --- | --- | --- |
| STN Confidence | $2.31 \pm 0.34$ | $2.60 \pm 0.38$ | $2.83 \pm 0.20$ |
| MRG Confidence | $2.95 \pm 0.14$ | $2.80 \pm 0.22$ | $2.98 \pm 0.08$ |
| STN Accuracy | 0.82 [0.75, 0.90] | 0.81 [0.59, 1.00] | 0.99 [0.96, 1.00] |
| MRG Accuracy | 0.94 [0.91, 0.97] | 0.78 [0.57, 0.99] | 0.99 [0.97, 1.00] |
| STN FN Rate | 0.05 [0.01, 0.09] | 0.08 [0.00, 0.20] | 0.01 [0.00, 0.04] |
| MRG FN Rate | 0.00 [0.00, 0.00] | 0.08 [0.00, 0.20] | 0.01 [0.00, 0.03] |

**Table 7:** Accuracy and False Negative Rate for STN and MRG stratified by z-score. Reader Confidence (not included in Fig. 6) listed as mean*±*sd, Accuracy (Fig. 6F) and False Negative Rate (Fig. 6H) listed as mean [95% CI] across three Centiloid bins for STN and MRG processing.

| Z-score | < 1 | 1–3 | 3+ |
| --- | --- | --- | --- |
| STN Confidence | $2.31 \pm 0.34$ | $2.64 \pm 0.32$ | $2.76 \pm 0.34$ |
| MRG Confidence | $2.94 \pm 0.15$ | $2.81 \pm 0.23$ | $2.92 \pm 0.15$ |
| STN Accuracy | 0.81 [0.74, 0.89] | 0.94 [0.87, 1.00] | 0.98 [0.96, 1.00] |
| MRG Accuracy | 0.93 [0.89, 0.97] | 0.91 [0.86, 0.95] | 0.99 [0.98, 1.00] |
| STN FN Rate | 0.06 [0.02, 0.09] | 0.03 [0.00, 0.07] | 0.02 [0.00, 0.04] |
| MRG FN Rate | 0.01 [0.00, 0.02] | 0.05 [0.00, 0.09] | 0.01 [0.00, 0.02] |

## 4 Discussion

This study demonstrates that MRG improves amyloid PET interpretability while preserving quantitative measures used for standardized clinical and research assessment. Across multiple amyloid tracers, MRG preserved SUVr and Centiloid metrics (Fig. 3), improved concordance between visual interpretation and quantitative burden measures (Fig. 4), increased radiologistrated image quality and confidence, and improved reader performance metrics relative to STN images (Fig. 5). The greatest benefit was observed in cases with low amyloid burden (Fig. 6, Supp. Fig. 2), which are among the most difficult amyloid PET scans to interpret accurately and confidently in practice [6, 10, 13, 14]. Together, these findings suggest that MRG is not only an image enhancement method, but also a practical strategy for improving amyloid PET readability where interpretation is most fragile.

Low amyloid burden cases are the most vulnerable to interpretive error [35] and are among the cases where early identification is most clinically relevant in the era of anti-amyloid therapy [4–6, 8, 9]. Early or borderline amyloid PET scans often show only subtle cortical uptake above adjacent white matter [10, 14, 19], requiring readers to rely on small regional differences rather than obvious diffuse cortical binding to make a diagnosis [18, 19, 22]. At conventional PET resolution, blur, spill-in, and partial volume effects can obscure the gray-white boundary and make early cortical changes due to AD difficult to detect [15, 17–19]. Burden-stratified analyses (Fig. 6) showed that MRG improved reader confidence across all severity ratings, while stratifying accuracy by severity score did not show statistical difference between MRG and STN.

However, accuracy clearly improved with MRG in low-binned Centiloid and z-scores. This discrepancy may be a result of the fact that sparse reader severity ratings for both MRG and STN were only associated with majority-negative amyloid reads, while low-binned Centiloid and z-scores capture both negative and borderline amyloid positive cases. Raters classified more amyloid negative cases as sparse with MRG as compared to STN (MRG: *n* = 61, STN: *n* = 41), suggesting improved visual identification of cases without appreciable cortical amyloid uptake in addition to improved visualization of low-burden amyloid cases. This shift in the number of cases classified as sparse with MRG may also explain the difference in Centiloid scores between STN and MRG for mild severity. This interpretation is further supported by the improved concordance between reader positivity and quantitative amyloid burden with MRG (Fig. 4). MRG increased AUC between visual reads and both Centiloid and z-score measures, suggesting that the enhanced images brought reader classifications into closer alignment with quantitative amyloid burden rather than simply shifting reads in one direction. Together, these results suggest that MRG has its strongest effect in low-burden and borderline amyloid PET cases, where improved cortical definition may reduce visual-quantitative discordance and support more confident interpretation near the threshold of visual positivity and negativity.

Our findings suggest that much of the mismatch between visual interpretation and quantification in amyloid PET reflects limitations of image interpretability due to poor image quality that can be improved upon with MRG denoising and resolution enhancement. Prior studies have shown that quantitative measures such as Centiloid can support visual reads, especially in challenging or borderline cases [10, 11, 19]. Even so, readers and quantitative burden measures may still disagree in practice [8, 10, 19]. In this study, MRG increased ROC/AUC concordance between reader positivity with both Centiloid and z-score measures relative to STN (Fig. 4). Therefore, MRG facilitates harmonization between visual interpretation and quSUPantitative amyloid burden. Furthermore, because MRG preserved SUVr and Centiloid quantification while improving this concordance, these findings suggest that improving image interpretability can reduce visual-quantitative mismatch without sacrificing the existing quantitative framework used to support amyloid PET assessment.

By evaluating MRG across essentially all clinically relevant amyloid PET tracers, this study moves MRG beyond a single-tracer demonstration and towards broader clinical applicability. Across FBP, FBB, FMM, and PiB, MRG consistently preserved quantitative behavior and showed that its reader-facing benefits were not confined to a single tracer or specialized acquisition settings. Across all tracers, the largest benefits were observed in low-burden cases, supporting MRG as a targeted strategy for improving amyloid PET interpretation where it is most challenging.

Recent complementary work by Khalighi et al. demonstrated improved reader agreement and preserved Centiloid behavior for FBB using an MR-guided PET reconstruction approach developed for simultaneous PET/MR imaging [28]. Our study is consistent with the findings of Khalighi et al, showing that MRG improves reader agreement but further demonstrates that it improves the confidence and accuracy of radiologist reads as well. A major difference is the significant acquisition and scanner heterogeneity of our datasets. Our study included 33 and 24 distinct PET and MRI scanner types, while the MRI and PET study dates for individual subjects ranged between same-day acquisitions to scans separated by up to 3.7 years. These findings suggest that MRG can be implemented in a vendor-neutral post-reconstruction optimization framework without requiring dedicated simultaneous PET/MR hardware. This supports the approach as a more scalable route than replacing installed scanners with newer high-performance systems. MRG may therefore represent a practical route for extending the benefits of anatomically informed PET denoising and resolution enhancement across the heterogeneous scanners and acquisition conditions that define real-world amyloid PET imaging.

This study has several limitations. Reader performance was evaluated against a leave-one-reader-out reference standard (based on 4 total readers) rather than an external ground truth, limiting interpretation of absolute diagnostic accuracy. Variable sample sizes across tracers and subgroup analyses reduced statistical power in some comparisons. Heterogeneous acquisition protocols across sites may have introduced variability in multi-tracer analyses. Both these sources of variability however are strengths regarding the anticipated external validity of the findings. Image review was performed using standardized slide decks rather than a clinical PACS environment to increase efficiency and consistency for time-intensive reader evaluations.

Future studies could evaluate MRG denoising and resolution enhancement prospectively in routine clinical workflows and determine its impact on reporting, confidence, and treatment-related decision making, particularly in cases of early or borderline amyloid positivity. Because current clinical visual-read training is based largely on standard-resolution images, future workflow studies should also determine whether formal reader training materials require updating for MRG images; however, the improved reader confidence, image quality, and performance observed here suggest that MRG may be readily interpretable by readers already trained on conventional amyloid PET. In addition, the performance of MRG is currently being investigated for Tau PET imaging to support a more comprehensive role in Alzheimer’s disease diagnosis and staging. Finally, direct comparisons with partial volume correction and other enhancement strategies, including high resolution PET systems [24], would further define the clinical role of this MRG approach.

In conclusion, MRG denoising and resolution enhancement improved perceived image quality, reader confidence, visual interpretation, and reader performance while preserving established quantitative amyloid PET behavior across tracers compared to STN. The greatest benefit was observed in low-burden scans near the threshold of visual positivity, where interpretation is most challenging and the clinical impact is the highest. These results support MRG post-processing as a practical and vendor-neutral strategy for improving amyloid PET interpretation while maintaining compatibility with current quantitative frame-works.

## Data Availability

The datasets analyzed in this study included publicly accessible research imaging datasets obtained through the Alzheimer's Disease Neuroimaging Initiative (ADNI/GAAIN) as well as retrospective de-identified clinical imaging datasets obtained under data-use agreements. Derived quantitative data supporting the findings of this study are available from the corresponding author upon reasonable request, subject to institutional and data-sharing restrictions.

https://adni.loni.usc.edu/

## Acknowledgements

Data used in preparation of this article were obtained from the Alzheimer’s Disease Neuroimaging Initiative (ADNI) database (https://adni.loni.usc.edu). As such, the investigators within the ADNI contributed to the design and implementation of ADNI and/or provided data but did not participate in the analysis or writing of this report. A complete listing of ADNI investigators can be found at: http://adni.loni.usc.edu/wp-content/uploads/how_to_apply/ADN_I_Acknowledgement_List.pdf.

Data collection and sharing for the Alzheimer’s Disease Neuroimaging Initiative (ADNI) is funded by the National Institute on Aging (National Institutes of Health Grant U19AG024904). The grantee organization is the Northern California Institute for Research and Education. In the past, ADNI has also received funding from the National Institute of Biomedical Imaging and Bioengineering, the Canadian Institutes of Health Research, and private sector contributions through the Foundation for the National Institutes of Health (FNIH), including contributions from AbbVie; Alzheimer’s Association; Alzheimer’s Drug Discovery Foundation; Araclon Biotech; BioClinica, Inc.; Biogen; Bristol-Myers Squibb Company; CereSpir, Inc.; Cogstate; Eisai Inc.; Elan Pharmaceuticals, Inc.; Eli Lilly and Company; EuroImmun; F. Hoffmann-La Roche Ltd and its affiliated company Genentech, Inc.; Fujirebio; GE Healthcare; IXICO Ltd.; Janssen Alzheimer Immunotherapy Research & Development, LLC; Johnson & Johnson Pharmaceutical Research & Development LLC; Lumosity; Lundbeck; Merck & Co., Inc.; Meso Scale Diagnostics, LLC; NeuroRx Research; Neurotrack Technologies; Novartis Pharmaceuticals Corporation; Pfizer Inc.; Piramal Imaging; Servier; Takeda Pharmaceutical Company; and Transition Therapeutics.

We also acknowledge the Alzheimer’s Association and the Global Alzheimer’s Association Interactive Network (GAAIN) for enabling access to datasets and reference frameworks used in this research.

## Supplementary Information

**Table S1:** Unique PET and MRI scanner types used in GAAIN datasets for the four amyloid tracers.

| Tracer | PET Scanners | MRI Scanners |
| --- | --- | --- |
| FBP | Siemens ECAT Exact HR+; Philips Gemini TF TOF 64; GE Advance | Not recoverable from local headers |
| FBB | Philips Medical Systems Gemini TF TOF 64 | Siemens TrioTim; Siemens MAGNETOM Skyra; GE Medical Systems Signa HDxt; Philips Ingenia; Siemens Aera; Siemens Avanto |
| FMM | Siemens Biograph 16 PET/CT; Siemens ECAT Exact HR+; GE Advance; GE Discovery 690 | GE Medical Systems Discovery MR750; GE Medical Systems Signa HDxt; Philips Medical Systems Achieva; Philips Medical Systems Intera; Siemens Trio; Siemens Allegra |
| PiB | Siemens Biograph TruePoint TrueV (Model 1094); Siemens ECAT Exact HR+; Siemens ECAT Exact HR; Philips Allegro PET scanner | Siemens TrioTim; GE Medical Systems SIGNA |

**Table S2:**
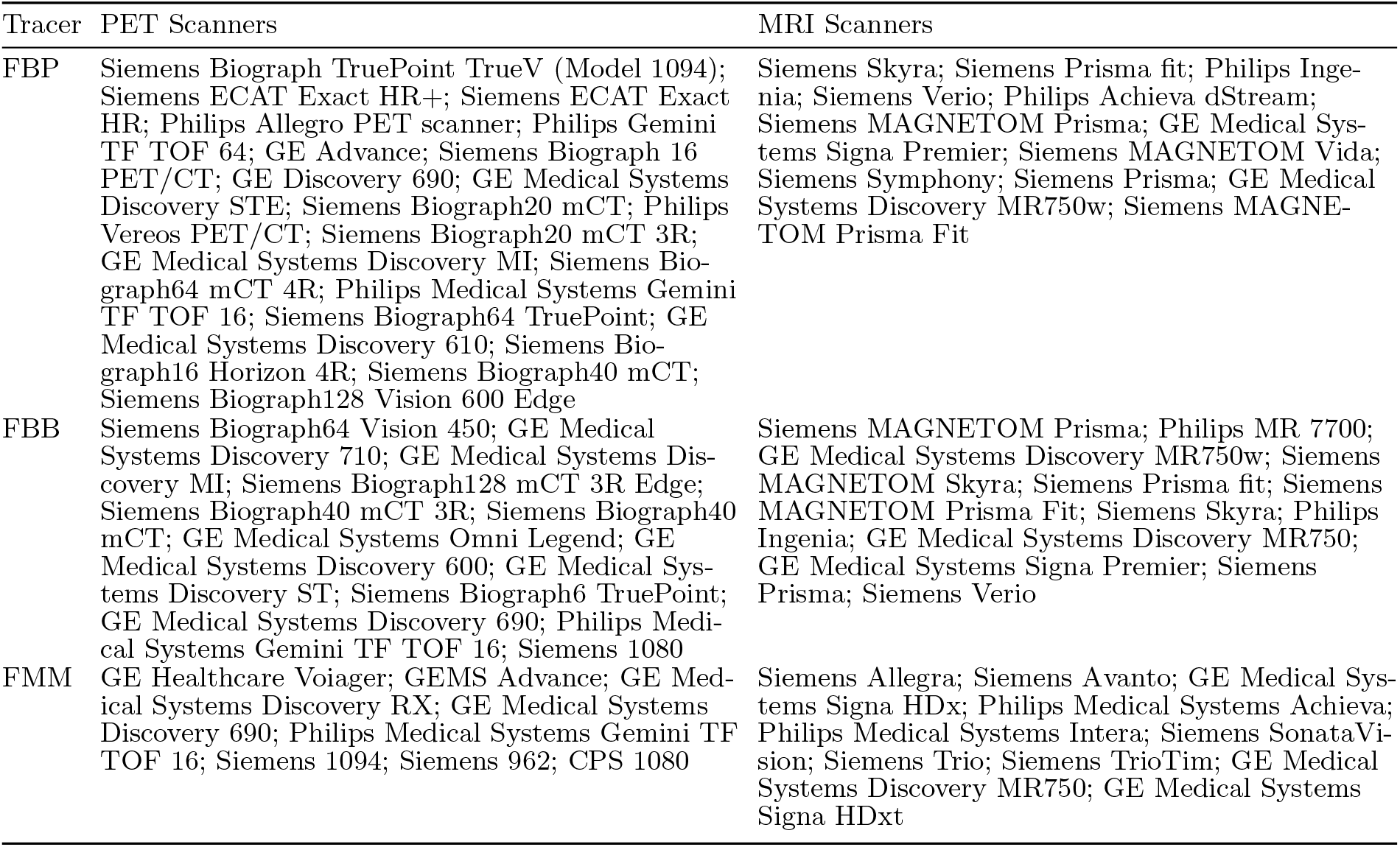
Unique PET and MRI scanner types used in the reader-study datasets for the three amyloid tracers.

**Fig. S1:**
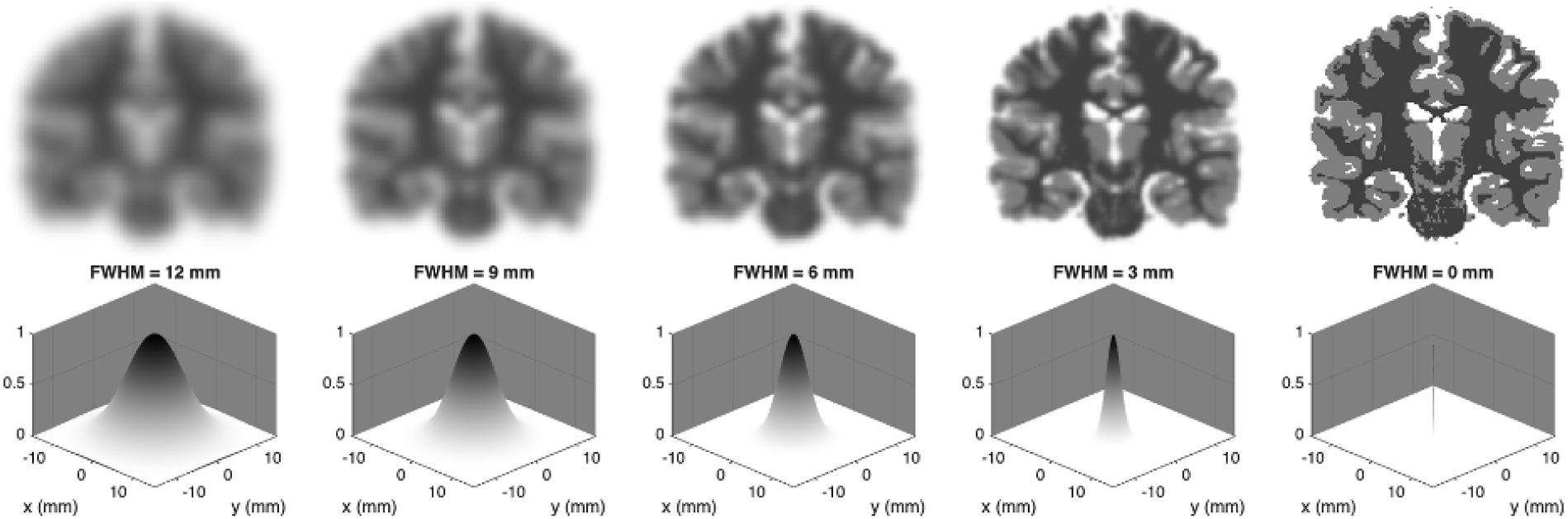
Simulation illustrating the image-contrast loss caused by partial-volume effect from PET blur and provides the conceptual basis for MRG. A BrainWeb PET phantom was convolved with Gaussian point-spread functions (FWHM = 12, 9, 6, 3, and 0 mm; left to right). The top row shows the point spread function kernels and the bottom row shows the resulting images. As the point spread function widens, the apparent contrast of PET images change. Broader point spread functions directly reduce the sharpness of the resulting PET images, while narrow functions enhance the image sharpness.

**Fig. S2:**
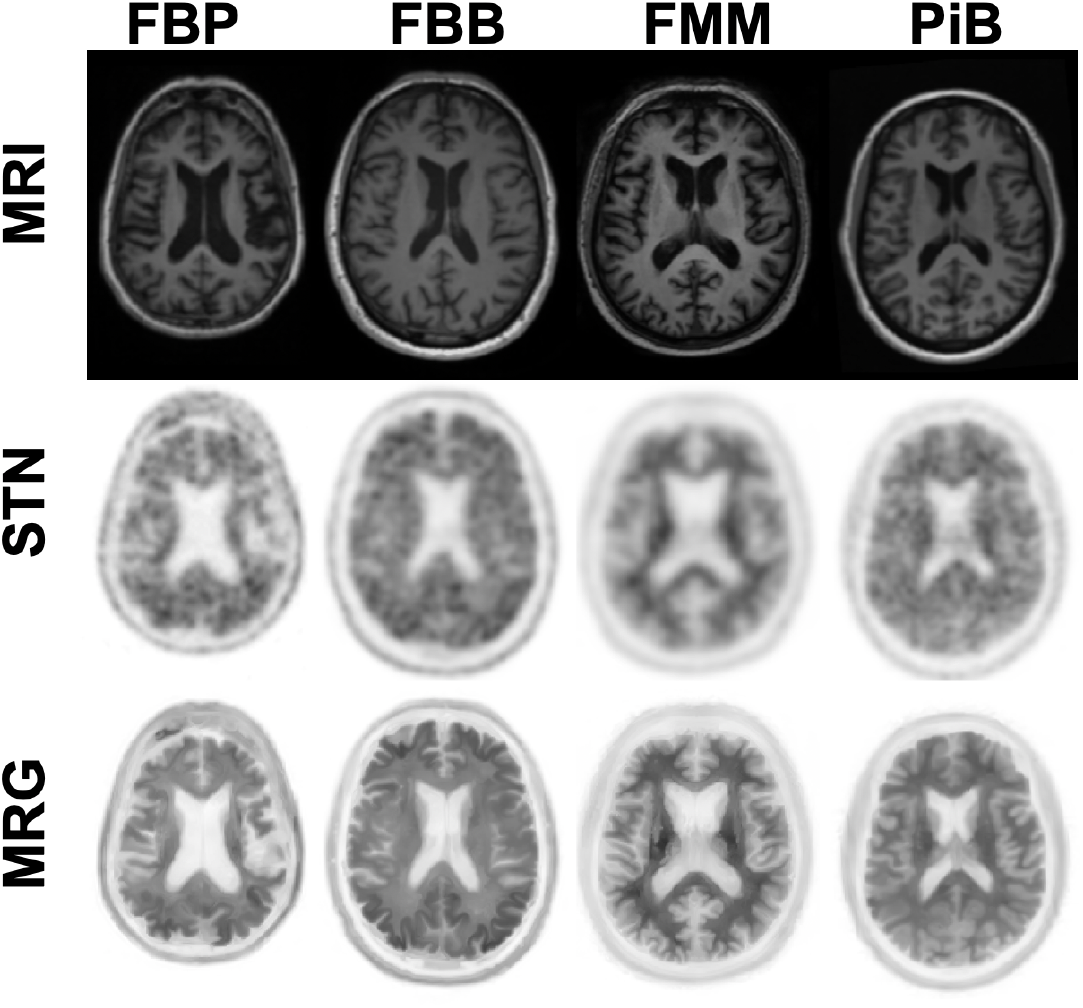
STN PET and MRG PET across tracers for low amyloid burden cases. Structural T1-weighted MRI is used to guide denoising and resolution enhancement of the standard PET reconstruction (STN), producing the MRI-guided PET output (MRG) for Florbetapir (FBP), Florbetaben (FBB), Flutemetamol (FMM), and Pittsburgh Compound B (PiB). Centiloids: FBP: 42, FBB: 36, FMM: 37, PiB: 24.

